# An empirical analysis of lay media coverage on influenza prevention pre- and post-COVID 19: Mask recommendations were previously rare, now ubiquitous

**DOI:** 10.1101/2023.02.14.23285818

**Authors:** Vinay Prasad, Elissa Brown, Alyson Haslam

## Abstract

**Importance:** Consistent, evidence-based communication is critical to building trust and maintaining credibility of public health agencies.

**Objective:** To identify any significant changes in the mainstream media’s presentation of public health advice for flu prevention before and after the COVID-19 pandemic.

**Design, Setting, and Participants:** A systematic search in Factiva of top ten U.S. newspapers by circulation, using two search periods, 2018-2019 and 2021-2022. Articles with flu prevention advice were identified, abstracted for media outlet, reporter, date. Articles were coded for the specific advice provided.

**Main Measure(s):** Number of recommendations for flu prevention, frequency of each recommendation; percent of recommendations aligned with CDC guidelines for each period. Changes in frequency of each recommendation. Differences determined using 2-proportion Z-tests, p-value 0.05 significance.

**Results:** 128 articles with 244 recommendations for pre-COVID period; 122 articles with 296 recommendations post-COVID. 96.3% of recommendations in alignment with CDC guidelines pre-COVID. 63.9% of recommendations in alignment with CDC during post-COVID timeframe. Percentage of articles with advice to mask for flu increased by 1,494.8% (p=<0.00001). 14.5% decline in percentage of articles advising flu vaccine (p=0.002). 495.5% increase in percentage of articles recommending social distancing (p=0.001). 1,368.9% increase in percentage articles recommending increased ventilation (p=0.0004).Advice to cover cough/sneeze declined by 52.8% (p=0.041); advice to disinfect surfaces declined by 76.7% (p=0.038).

**Conclusions and Relevance:** Expert advice on flu prevention as presented in top 10 U.S. newspapers changed significantly during the COVID-19 pandemic. The strategies discussed more frequently are not currently recommended by CDC. This is relevant information for public health leaders as they address ongoing issues of trust and credibility.

**Key Points:** *Question:* Are there significant differences between the mainstream media’s presentation of expert advice for flu prevention before and after the COVID-19 pandemic?

*Findings:* A systematic search of the top ten U.S. newspapers by circulation found that the percentage of total articles with advice to use a face mask for flu prevention increased by 1,494.8% from the pre-to post-COVID period, while the percentage of total articles advising a flu vaccine decreased by 14.5%. Other significant findings include an increase in advice to social distance (494.5%), an increase in advice to improve ventilation (1,368.9%), a decline in advice to cover your cough (58.2%) and a decline in advice to disinfect surfaces (76.7%). The strategies discussed more frequently are not currently recommended by Centers for Disease Control (CDC) for flu prevention.

*Meaning:* Significant changes in public health advice on flu prevention, as presented by high-circulation U.S. newspapers, occurred during the COVID-19 pandemic, resulting in less consistency with CDC recommendations for flu prevention.

## Introduction

Media outlets in the United States report on seasonal respiratory viruses each year, particularly in the fall and winter months. A portion of these articles feature advice on how to avoid contracting and spreading seasonal influenza as well as various treatment options if you are infected. The articles typically include one main call to action: get a flu vaccine.

Since 2010, the Centers for Disease Control and Prevention (CDC) has recommended the annual flu vaccine for everyone 6 months and older with few exceptions.^1^ Practices like frequent hand-washing, avoiding people who are sick, disinfecting surfaces, covering your mouth when you cough or sneeze, staying home when sick, and seeking treatment with antivirals are also commonly included in the lay media coverage, in line with CDC guidelines.

Hygiene-based flu prevention strategies were recommended as a means of controlling the spread of SARS-CoV-2, beginning in early 2020. When the World Health Organization (WHO) suggested countries take “urgent and aggressive action”, additional containment and mitigation measures were proposed by medical professionals and public health leaders in the United States and around the world.^2^ People were encouraged to avoid crowds, seek ways to improve ventilation such as opening windows, wash or wipe down groceries, and stand six feet apart (a practice known as “social distancing”). Plexiglass barriers were installed in public spaces and international travel restrictions were put in place.

On July 14, 2020 the CDC also called upon people living in the United States to wear face masks as a measure that could possibly reduce the spread of SARS-CoV-2.^3^ Information on the effectiveness of these control measures was derived almost entirely from observational or modeling studies, yet they were considered necessary in the face of a looming pandemic.

Scientists and medical providers have speculated about the role these various COVID-19 mitigation strategies – specifically masking, social distancing, and ventilation – played in reducing levels of flu and other seasonal respiratory viruses during the COVID-19 pandemic. The evidence to support community masking for flu prevention prior to the pandemic was sparse. In fact, a Cochrane Library meta-analysis on the effectiveness of physical interventions to interrupt or reduce the spread of acute respiratory viruses did not show a clear reduction in respiratory viral infection with the use of medical/surgical masks during flu season.^5^ Perhaps this is why masking is not currently recommended by the CDC for preventing influenza or RSV.^6,7^ With regard to social distancing, the Cochrane meta-analysis concludes that there is a need for large, well-designed RCTs in multiple settings and populations, as quality data does not exist to make a determination. There are currently no published RCTs on ventilation as a method of reducing the transmission of viruses.

In the absence of evidence from randomized control trials (RCTs), many experts have nevertheless began recommending masking, social distancing, and improved ventilation for the prevention of all seasonal respiratory viruses. These sentiments are now frequently presented as best practices for flu prevention in the news media, with a specific emphasis on masking. For example, Dr. John Swartzberg, an infectious disease expert at the University of California Berkeley School of Public Health, told Reuters in December 2022, “If you don’t want to get sick and you don’t want to go to the hospital and you don’t want to die of COVID or influenza, or if a very young child, RSV, then you should be wearing a mask indoors in a public place.”^4^

It is with this background that we sought to evaluate the potential influence of COVID-19 mitigation strategies on the media’s presentation of advice on preventing seasonal influenza, by reviewing articles on flu prevention published by high-circulation newspapers before and after the start of the COVID-19 pandemic.

## Methods

We intended to compare news articles providing influenza prevention and treatment advice before and after the COVID-19 pandemic to observe whether any significant changes have occurred.

### Data collection

We conducted a systematic search using Factiva, an archive of over 32,000 major global newspapers, newswires, industry publications, magazines, reports, and other sources. We limited the search to the print editions of the top ten newspapers in the United States by circulation to avoid duplications. These consist of The Wall Street Journal, The New York Times, USA Today, The Washington Post, The New York Post, The Los Angeles Times, The Chicago Tribune, The Minneapolis Star Tribune, Newsday, and The Boston Globe.

We included two search periods: 1) January 1, 2018 through December 31, 2019, which represented the pre-pandemic time period, and 2) January 1, 2021 to December 31, 2022, which represented the post-pandemic time period. We omitted 2020, as this year marked widespread global concern with COVID-19 and shifting recommendations; it is difficult to select a single date to disambiguate a pre- and post-COVID-19 time period.

The search terms we used in Factiva were: ‘flu’ or ‘influenza’ and ‘prevent’; ‘flu or influenza’ and ‘avoid’, ‘flu or influenza’ and ‘recommend’. The search was inclusive of all types of print newspaper articles including news reports, feature articles, editorials, columns and opinion (op-ed). Articles were selected for analysis if they included a call to action for flu prevention or treatment, such advice from public health entities (CDC, state and local health departments, government officials), medical professionals, scientists, or directly from the reporter.

These articles were then coded for 11 recommendations. Eight of the 11 were based on CDC guidelines from 2018-2022, and include: flu vaccination, handwashing, take antivirals if prescribed, stay home or isolate when sick, cover your cough or sneeze, disinfect surfaces, avoid sick people, and don’t touch your face (See Table 1). We coded for three additional measures that have become more common in the lay media since the COVID-19 pandemic: wear a mask, practice social distancing, and improve ventilation. For each article, we abstracted the following information: media outlet, reporter, date, and the advice or recommendations being advocated.

**Table 1:**
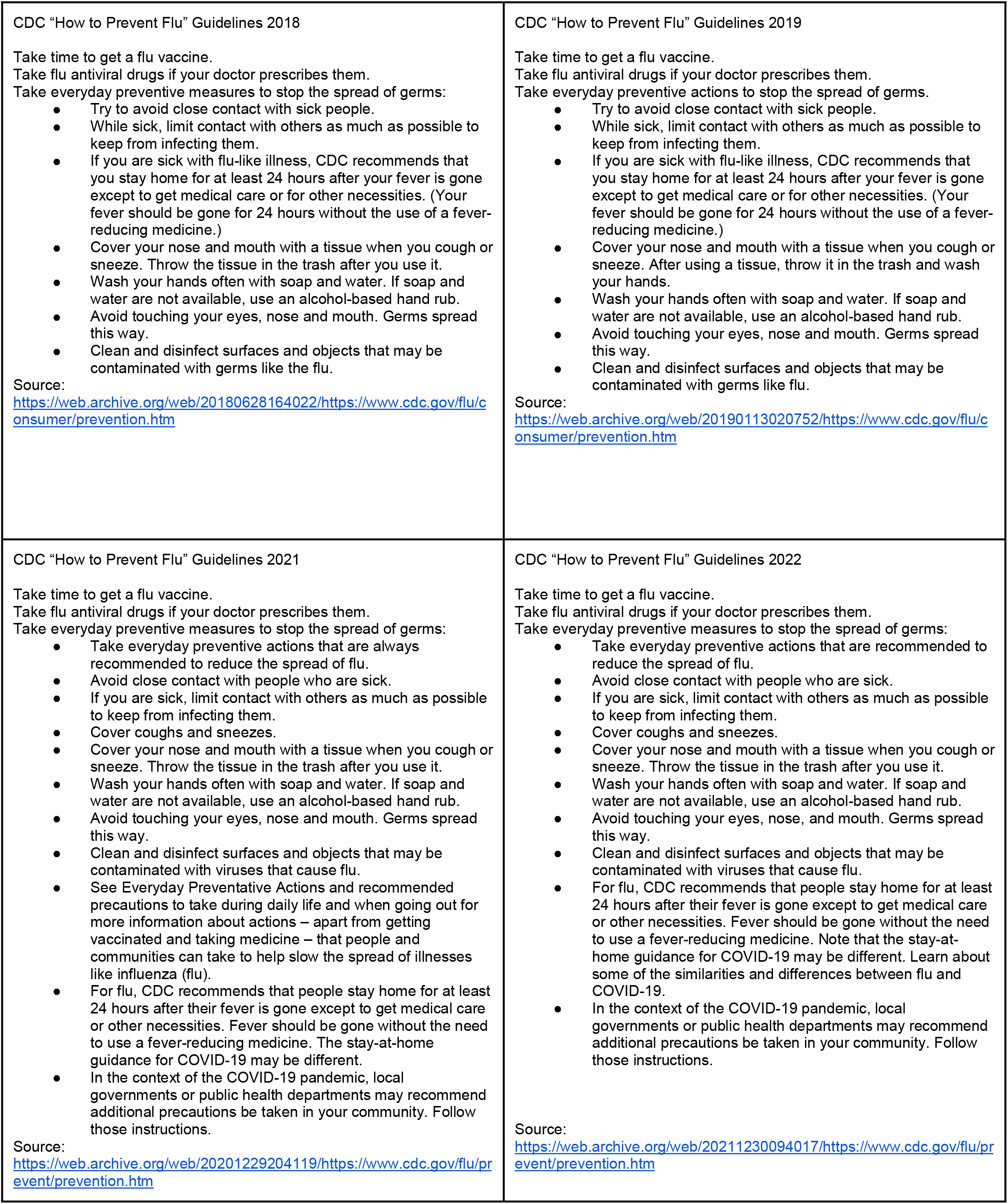
CDC Flu Prevention Guidelines, 2018-2022.

**Table 2:** Comparison of flu prevention and treatment recommendations in top 10 U.S. newspapers before and after the COVID-19 pandemic.

| Guideline | Articles |  |  |  | Recommendations |  |  |  |
| --- | --- | --- | --- | --- | --- | --- | --- | --- |
|  | % of total 2018-2019 (n=128) | % of total 2021-2022 (n=122) | Relative change | p-value | % of total 2018-2019 (n=244) | % of total 2021-2022 (n=296) | Relative change | p-value |
| Flu vaccine | 93.0% | 79.5% | -14.5% | **0.002** | 48.8% | 32.8% | -32.8% | **0.0002** |
| Wash hands | 22.7% | 24.6% | 8.5% | 0.719 | 11.9% | 10.1% | -14.7% | 0.516 |
| Antivirals | 17.2% | 13.1% | -23.7% | 0.368 | 9.0% | 5.4% | -40.0% | 0.103 |
| Stay home when sick | 17.2% | 23.0% | 33.5% | 0.254 | 9.0% | 9.5% | 4.9% | 0.857 |
| Cover cough | 15.6% | 7.4% | -52.8% | **0.041** | 8.2% | 3.0% | -62.9% | **0.008** |
| Disinfect surfaces | 7.0% | 1.6% | -76.7% | **0.038** | 3.7% | 0.7% | -81.7% | **0.014** |
| Avoid sick people | 5.5% | 3.3% | -40.0% | 0.401 | 2.9% | 1.4% | -52.9% | 0.215 |
| Don't touch face | 5.5% | 2.5% | -55.0% | 0.226 | 2.9% | 1.0% | -64.7% | 0.112 |
| Masks | 3.9% | 62.3% | 1494.8% | **< 0.00001** | 2.0% | 25.7% | 1153.0% | **< 0.00001** |
| Social distance | 2.3% | 13.9% | 494.5% | **0.001** | 1.2% | 5.7% | 367.1% | **0.006** |
| Ventilation | 0.8% | 11.5% | 1368.9% | **0.0004** | 0.4% | 4.7% | 1054.1% | **0.002** |

**Table 3:** Flu prevention and treatment articles from before and after the COVID-19 pandemic, segmented by media outlet.

| Ranking<sup>a</sup> | News Outlet | # articles 2018-2019 | % of total | # articles 2021-2022 | % of total | Relative change (%) | p-value |
| --- | --- | --- | --- | --- | --- | --- | --- |
| 1 | USA Today | 9 | 7.0% | 10 | 8.2% | 16.6% | 0.748 |
| 2 | The Wall Street Journal | 11 | 8.6% | 10 | 8.2% | -4.6% | 0.756 |
| 3 | New York Times | 27 | 21.1% | 28 | 23.0% | 8.8% | 0.849 |
| 4 | New York Post | 6 | 4.7% | 2 | 1.6% | -65.0% | **0.046** |
| 5 | Los Angeles Times | 13 | 10.2% | 13 | 10.7% | 4.9% | 1 |
| 6 | The Washington Post | 20 | 15.6% | 24 | 19.7% | 25.9% | 0.395 |
| 7 | Minneapolis Star-Tribune | 9 | 7.0% | 6 | 4.9% | -30.1% | 0.271 |
| 8 | Newsday | N/A | N/A | N/A | N/A | N/A | N/A |
| 9 | Chicago Tribune | 19 | 14.8% | 11 | 9.0% | -39.3% | **0.038** |
| 10 | The Boston Globe | 14 | 10.9% | 18 | 14.8% | 34.9% | 0.317 |

<sup>a</sup>"Top 10 U.S. Daily Newspapers". Cision. January 4, 2019

Because we used publicly available data and did not use personal information, we were not required to obtain institutional review board approval, in accordance with 45 CFR §46.102(f).

### Statistical analysis

First, for each time period, we determined the total number of recommendations for influenza prevention and calculated the frequency of each recommendation, both as a percentage of articles and as a percentage of total recommendations. We then determined what percent of the recommendations aligned with the guidelines on the CDC’s “How to Prevent Flu” web page for each time period.

Relative changes in the frequency of each recommendation as a percent of total articles and as a percent of total recommendations were calculated. We calculated differences in percentages, before and after COVID-19 with 2-proportion Z-tests, using a p-value of 0.05 as statistical significance. We used Social Science Statistics’ Z Score Calculator for 2 Population Proportions to conduct data analysis.

## Results

In total, 128 articles met our inclusion criteria for 2018-2019 and 122 articles for 2021-2022. 244 recommendations for flu prevention and treatment were coded from the pre-COVID time period; 296 recommendations for flu prevention and treatment were coded from the post-COVID time period.

During 2018-2019, 93.0% (119/128) of all articles in the analytic sample recommended getting a flu vaccine, 22.7% (29/128) recommended washing hands, 17.2% (22/128) recommend seeking antivirals, 17.2% (22/128) recommended staying home when sick, 15.6% (20/128) recommended covering your cough or sneeze, 7.0% (9/128) recommended disinfecting surfaces, 5.5% (7/128) recommend avoiding sick people, 5.5% (7/128) recommend not touching face, nose or eyes, 3.9% (5/128) recommended face masks, 2.3% (3/128) recommend social distancing, and 0.8% (1/128) recommended improved ventilation (see Figure 1). 96.3% of these recommendations were in alignment with CDC guidelines from 2018-2019 for preventing and treating influenza.

**Figure 1:**
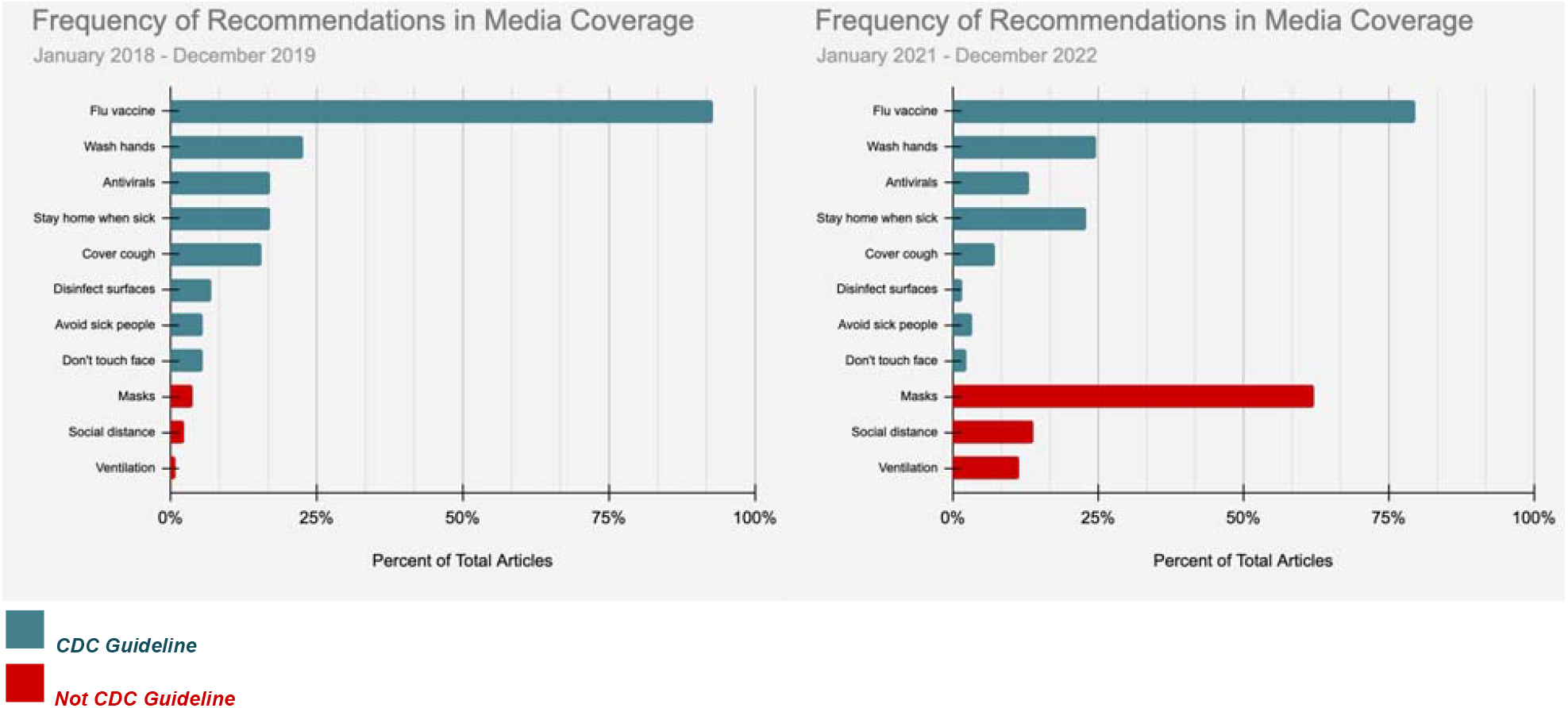
Frequency of flu prevention and treatment advice in the media before and after the COVID-19 pandemic.

During 2021-2022, 79.5% (97/121) of all articles in the analytic sample recommended getting a flu vaccine, 62.3% (76/121) recommend wearing a face mask, 24.6% (30/121) recommended washing hands, 23.0% (28/121) recommended staying home when sick, 13.9% (17/121) recommend social distancing, 13.1% (16/121) recommend seeking antivirals, 11.5% (14/121) recommend improved ventilation, 7.4% (9/121) recommend covering your face when you cough or sneeze, 3.3% (4/121) recommend avoiding sick people, 2.5% (3/121) recommend not touching your face, nose or eyes, 1.6% (2/121) recommend disinfecting surfaces (See Figure 1). 63.9% of these recommendations were in alignment with CDC guidelines from 2021-2022 for preventing and treating influenza.

We found that the percentage of articles with advice to use a face mask for influenza prevention was significantly higher in 2021-2022 than in 2018-2019 (3.9% vs. 62.3%; p=<0.00001), an increase of 1,494.8%. Masking as a percent of total recommendations for flu prevention increased by 1,153.0% from 2.0% to 25.7% (p=<0.00001). There was a 14.5% decline in the percentage of articles advising a flu vaccine (p=0.002) and a decline of 32.8% in recommendations for flu vaccines as a percent of total recommendations (p=0.0002).

Other large differences include a 495.5% increase in the percentage of articles recommending social distancing, from 2.3% to 13.9% (p=0.001). A 1,368.9% increase in the percentage articles that recommended increased ventilation, from 0.8% to 11.5% of articles (p=0.0004). The percent of articles with advice to cover your cough or sneeze declined by 52.8% from 15.6% of articles to 7.4% of articles (p=0.041), while the percent of articles with advice to disinfect surfaces declined by 76.7% (p=0.038) from 7.0% to 1.6% of articles (see Figure 2).

**Figure 2:**
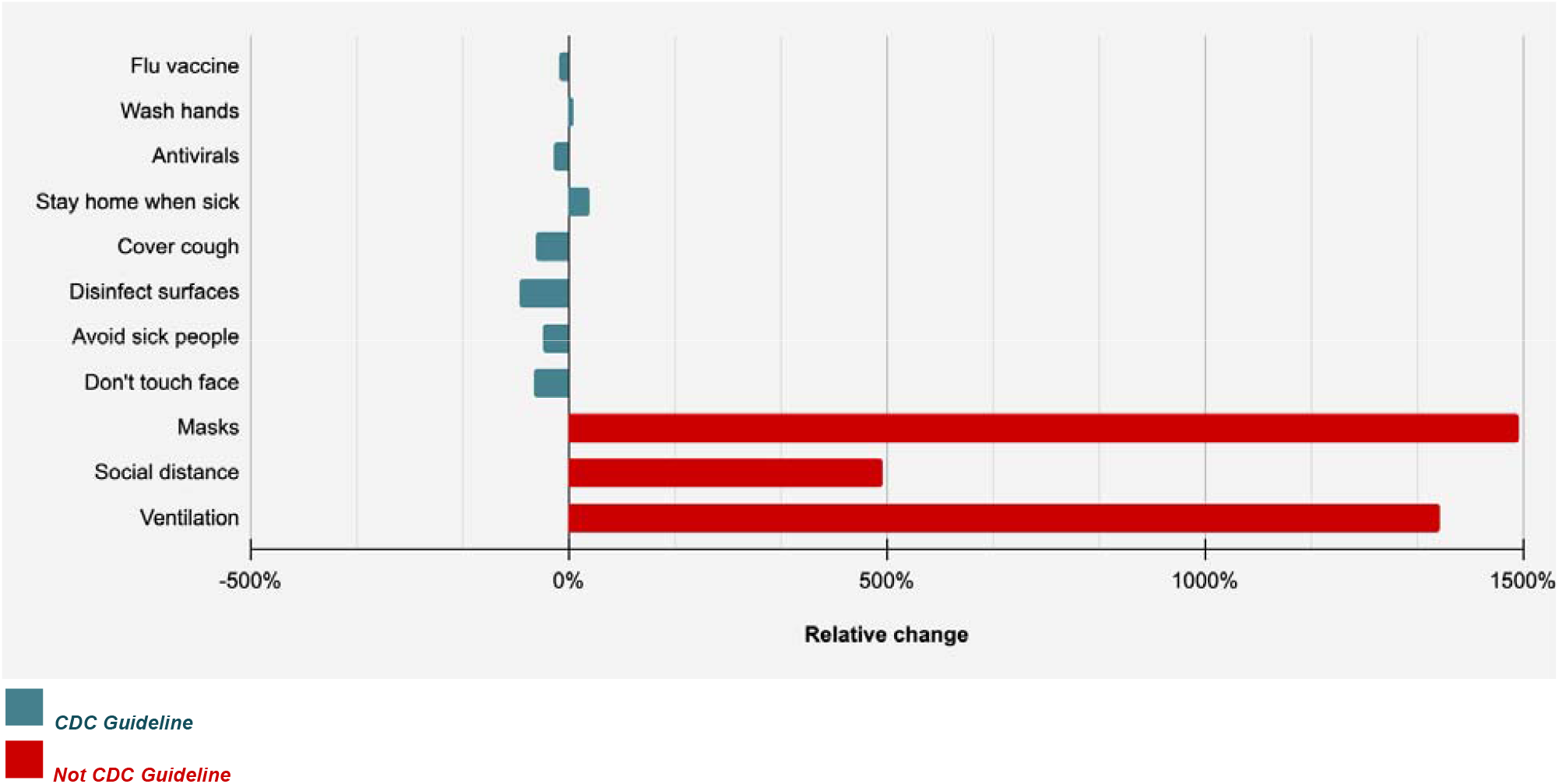
Relative change in flu prevention and treatment advice in the media before and after the COVID-19 pandemic.

No significant differences were observed in the percent of articles recommending hand washing, seeking antivirals, staying home when sick, avoiding sick people, and not touching your face, nose, and eyes.

Our analysis found a 42.1% decrease in the number of articles written by The Chicago Tribune, from 19 articles in 2018-2019 to 11 articles in 2021-2022 (p= 0.038). The number of article on flu prevention also declined in the New York Post by 66.7% (p=0.046), but the total number of article was small (n=8) so these findings should be interpreted with caution. There were no significant changes identified in the number of articles on flu prevention in The Wall Street Journal, The New York Times, USA Today, The Washington Post, The Los Angeles Times, The Minneapolis Star Tribune, and The Boston Globe.

## Discussion

In our review of high-circulation U.S. newspapers, we found differences in the advice presented for the prevention of seasonal influenza in the years before the COVID-19 pandemic and in the years after the start of the COVID-19 pandemic. Most notably, strategies like getting a flu vaccine, covering one’s mouth during a sneeze or cough, and disinfecting surfaces are advised less frequently, while strategies like wearing face masks, social distancing, and improvements in ventilation are discussed more frequently. Moreover, the strategies that are discussed more frequently are not ones that are currently recommended by the CDC for flu prevention.

These changes in advice for flu prevention occurred during a period of time without a dramatic change in the evidence base for flu prevention. The Cochrane Library recently published an updated meta-analysis on the use of physical interventions to interrupt or reduce the spread of respiratory viruses, which reports minimal changes to the evidence from the pre-COVID to post-COVID period. Evidence for masking – specifically for influenza prevention – remains “poor.” The Cochrane meta-analysis includes RCTs and cluster RCTs. Combining 78 trials and 276,917 participants, the analysis concludes that wearing masks in the community probably makes little or no difference to the outcome of influenza-like or COVID-19-like illness compared to not wearing masks (RR 0.95, 95% CI 0.84 to 1.09)8. Regarding social distancing, the authors maintain that there is a need for large, well-designed RCTs in multiple settings and populations, as quality data does not exist to make a determination. There are still no completed RCTs on ventilation as a method of reducing the transmission of respiratory viruses.

The public is increasingly skeptical about the credibility of public health information. In fact, trust in scientists and medical providers declined over the course of the COVID-19 pandemic and is now below pre-pandemic levels.9 Issues of trust stemming from the pandemic have the potential to “spill over” and impact the success of other public health efforts. For example, polarization surrounding the COVID-19 vaccine appears to have created vulnerabilities with regard to annual flu vaccination efforts. Specifically low Covid-19 vaccination rates were found to be associated with decreases in influenza vaccination rates.10 Further, 31.8% of respondents to a recent CDC survey indicated that they “probably” or “definitely will not” receive a flu vaccine this year.11

Evidence of declining enthusiasm toward childhood vaccination campaigns in the United States is also concerning. More than half of parents in a recent poll say they “definitely won’t” get their child under five vaccinated against COVID-19.12 Opposition to requiring the childhood measles, mumps and rubella (MMR) vaccine now stands at 35%, up from 23% in 2019, and the share of the general public that says healthy children should be required to get those vaccines to attend public schools is down 11% from 2019.13 Finally, coverage against measles in the U.S. dropped to the lowest it’s been in more than a decade.14

Misinformation campaigns and the expanding influence of social media are commonly blamed for the erosion of trust in public health and resulting “vaccine hesitancy”. However, it is important to consider the impact of rapidly shifting messages from public health leadership, scientists, and healthcare experts in the absence of supporting evidence. The public was introduced to many new interventions aimed at reducing the spread of COVID-19 in 2020, including masking, social distancing, and increased ventilation. These strategies are not included in the CDC’s annual flu prevention guidelines and were only rarely recommended by health experts in the news media before the pandemic. Trust begins with accountability, yet dramatic shifts in expert recommendations have occurred without changes in the available evidence.

### Limitations

There are several limitations to our analysis. First, we used the top ten print newspapers in the United States by circulation, so our findings may not apply to the lay media at-large. Second, we only used the two years prior to and two years after the introduction of COVID-19, which may limit generalizability to other years. Third, our search terms may not have captured all articles in each news source on the topic at hand, but we did use a systematic search, so our results are likely not differentially affected. Finally, articles from Newsday were not available through our research database.

## Conclusions

The role of the mainstream media as a vehicle for credible public health communication is especially important at a time when people in the United States are increasingly turning to less credible sources – such as social media – for health information. The endorsement of preventive measures that are without efficacy data and that are unsupported by federal recommendations may detract attention from other well-established preventive measures. Medical and public health experts may want to consider the benefits of communicating consistent, evidence-based recommendations through the media as a means of regaining trust and building credibility.

## Data Availability

All data produced in the present study are available upon reasonable request to the authors

